# Interactions between nuclear and mitochondrial SNPs and Parkinson’s disease risk

**DOI:** 10.1101/2022.01.24.22269732

**Authors:** Sarah J. Pickett, Dasha Deen, Angela Pyle, Mauro Santibanez-Koref, Gavin Hudson

**Affiliations:** Clinical and Translational Sciences Institute, Faculty of Medical Sciences, Newcastle University, Newcastle upon Tyne, NE2 4HH, UK; Biosciences Institute, Faculty of Medical Sciences, Newcastle University, Newcastle upon Tyne, NE2 4HH, UK

**Keywords:** Case only design, Parkinson’s disease, Mitonuclear combinations

## Abstract

Interactions between the products of the nuclear and mitochondrial genomes are critical for the function of most eukaryotic cells. Recently the introduction of mitochondrial replacement therapy has raised the question of incompatibilities between mitochondrial and nuclear variants, and their potential influence on the genetic makeup of human populations. Such interactions could also contribute to the variability of the penetrance of pathogenic DNA variants. This led us to investigate the frequencies of combinations of nuclear and mitochondrial SNP alleles (mitonuclear combinations) in healthy individuals (n=5375) and in a cohort of patients with Parkinson’s disease (PD, n=2210). In the unaffected population, we were not able to find associations between nuclear and mitochondrial variants with a false discovery rate below 0.05 after accounting for multiple testing (i.e., the number of combinations examined). However, in the PD cohort, five combinations surpassed this threshold. Next, combining both cohorts, we investigated whether these associations were modulated by disease status. All five combinations were significant (p<10^−3^ for all tests).These combinations also showed significant evidence for an effect of the interaction between the mitochondrial and nuclear variants on disease risk. Their nuclear components mapped to *TBCA, NIBAN3* and *GLT25D1* and an uncharacterised intergenic region. In summary, starting from a single cohort design we identified combinations of nuclear and mitochondrial variants affecting PD disease risk.

## Introduction

The physiological function of eukaryotic cells requires contributions from both mitochondrial and mitochondrial gene products [1]. Sequence variation in the nuclear (nDNA) or mitochondrial (mtDNA) genome can lead to phenotypic changes [1]. Diseases linked to specific mtDNA variants, such as mt.3243 or mt.11778 [1], show wide differences in penetrance [2, 3] and studies indicate that the nDNA contributes to these differences [3, 4].

Common mtDNA variants have been repeatedly associated with complex disease, particularly with diseases involving neurological degeneration [5-11]. However, beyond a few examples such as Parkinson’s disease (PD) [5-7, 12], such associations have proved difficult to replicate [13]. This could reflect the small effect sizes associated with mtDNA variants [5, 6] or modulation of their effects by nDNA variation. Recently the introduction of mitochondrial replacement therapy [14-16], has raised the question of whether mitonuclear incompatibilities exist [15, 17], and more generally, the extent to which these incompatibilities may influence health [18].

Within a population, the frequencies of combinations of nuclear and mitochondrial variants will reflect population structure as well as selection. In a cohort of patients affected by a specific disease, and where disease risk is influenced by the interaction from mitochondrial and nuclear variants, depletion or enrichment of corresponding combinations will be detectable as deviations from the frequency expected given the frequencies of the individual variants in the cohort. This has motivated using measures of association between alleles to detect such interactions (‘mitonuclear associations’). However, these associations could also reflect admixture or other forms of population stratification. This is of particular interest because mitochondrial variants have been extensively used to track the migration of populations and ancestry [19].

Mitonuclear associations have been repeatedly investigated [20-22]. An analysis of Human Genome Diversity Project data identified significant, albeit weak, mitonuclear associations through linkage disequilibrium [20]. However, recent work in the Japanese population found no evidence for such associations [21]. The importance of considering population structure was also highlighted by a recent analysis of whole-genome sequencing data that showed evidence of correlations between nuclear and mitochondrial alleles in specific geographic regions [22].

In this communication, we investigate mitonuclear associations using a GWAS approach with mitochondrial variants as the phenotype of interest. Using HapMap Phase 3 European sequence data, we selected variants with sufficient power to detect associations characterised by an odds ratio of at least 1.5 (**Supplementary Materials**). We selected two sets of European individuals genotyped on the same microarray system, one of patients with Parkinson’s disease (PD) and one of unselected individuals (detailed in **STable 1**). For each cohort, we performed a genome-wide search for nuclear polymorphisms showing significant association with each of the selected mitochondrial variants. After examining single cohorts, we expanded our analyses to incorporate disease status by assessing the interaction between disease status and nuclear variants on the association for the combinations that achieved significance in at least one of the two cohorts. Finally, we quantified the effect of the combinations that showed a significant result in the last step on disease risk, i.e., the association between disease stand and the interaction between mtSNP and nSNP.

## Results

First, we determined the minor allele frequency (MAF) of the mtSNP that would allow us to achieve a power of 0.8 to detect an association with an odds ratio of 1.5, for a nominal type-I error of 5×10^−8^, a nSNP with a MAF of 0.10 and a cohort size of 2000. This required the mtSNP to have a MAF>0.2. According to HapMap Phase 3 genome data, six mtSNPs met that threshold. However, three of these (mt.11467, mt.12308 and mt12372, **STable 2**) are strongly associated with each other (R^2^>0.9), with R^2^<0.5 for the remaining pairwise associations. Thus, we selected a subset of four mtSNPs (mt.2706, mt.3010, mt.11251 and mt.11467) for further analysis (**Supplementary Materials**).

Next, we assessed the associations between the nSNPs and the selected mtSNPs. For the unaffected individuals (cohort 58C/NBS; see **Supplementary Materials**) the Q-Q plots for all 4 mtSNPs are shown in **SFigure 1**. Taking this into consideration no mitonuclear associations achieved a false discovery rate (FDR) below 0.05. Conversely, for PD, five combinations achieved this threshold (**Table 1** and Q-Q plots in **SFigure 2** and locus plots in **SFigures 3-5**).

**Table 1.** Significant (FDR<0.05) mitonuclear combinations identified in the PD cohort. ^a^: Supplementary Figure reference; ^b^: Assessed using the term for the interaction between disease status and nSNP; ^c^: Assessed using the term for the interaction between mtSNP and nSNP; interg: Intergenic. Chromosome and position (chr:bp) are based upon GRC1137.

| Mitonuclear combination |  |  |  |  |  | Mitonuclear association |  |  |  |  |  | Association with PD |  |  |  |  |  |
| --- | --- | --- | --- | --- | --- | --- | --- | --- | --- | --- | --- | --- | --- | --- | --- | --- | --- |
| *mtSNP* |  |  | *nSNP* |  |  | in PD cohort |  |  |  | Effect of disease status on association<sup>b</sup> |  | mtSNP |  | nSNP |  | Mitonuclear combination<sup>c</sup> |  |
| *dbSNP ID* | *chr:bp* | *Locus* | *ID* | *chr:bp* | *Locus* | *P* | *FDR* | *OR*<br>(95% *CI*) | *Plot<sup>a</sup>* | *P* | *OR*<br>(95% *CI*) | *P* | *OR*<br>(95% *CI*) | *P* | *OR*<br>(95% *CI*) | *P* | *OR*<br>(95% *CI*) |
| rs2854128 | mt.2706 | *MT-RNR2* | rs11666267 | 19:17660300 | *NIBAN3* | 4.2E-08 | 1.5E-02 | 1.4<br>(1.3-1.6) | *SFig. 3* | 2.5E-07 | 1.5<br>(0.1-1.7) | 7.2E-02 | 0.6<br>(0.4-1.0) | 2.1E-01 | 1.1<br>(1.0-1.1) | 4.16E-06 | 1.5<br>(1.3-1.7) |
| rs2853493 | mt.11467 | *MT-ND4* | rs254412 | 5:77079452 | *TBCA* | 5.8E-09 | 1.0E-02 | 1.6<br>(1.4-1.8) | *SFig. 4* | 1.1E-05 | 1.5<br>(1.3-1.8) | 2.4E-01 | 0.9<br>(0.8-1.0) | 1.1E-03 | 1.1<br>(1.0-1.2) | 7.20E-05 | 1.5<br>(1.2-1.8) |
| rs2853493 | mt.11467 | *MT-ND4* | rs384109 | 5:77019909 | *TBCA* | 1.9E-08 | 1.2E-02 | 1.6<br>(1.3-1.8) | *SFig. 4* | 1.2E-05 | 1.5<br>(1.3-1.8) | 2.4E-01 | 0.9<br>(0.8-1.0) | 2.1E-03 | 1.1<br>(1.1-1.2) | 5.53E-05 | 1.5<br>(1.2-1.8) |
| rs2853493 | mt.11467 | *MT-ND4* | rs441492 | 5:76973352 | *TBCA* | 1.2E-08 | 1.1E-02 | 1.6<br>(1.3-1.8) | *SFig. 4* | 5.7E-05 | 1.4<br>(1.2-1.7) | 2.4E-01 | 0.9<br>(0.8-1.0) | 6.9E-03 | 1.1<br>(1.1-1.2) | 5.81E-05 | 1.5<br>(1.2-1.8) |
| rs2853493 | mt.11467 | *MT-ND4* | rs1606610 | 17:6301562 | *Interg* | 3.6E-08 | 1.5E-02 | 1.7<br>(1.4-2.0) | *SFig. 5* | 6.2E-06 | 1.7<br>(1.3-2.1) | 2.4E-01 | 0.9<br>(0.8-1.0) | 2.1E-01 | 1.1<br>(1.0-1.2) | 9.19E-07 | 1.9<br>(1.5-2.4) |

Next, we proceeded to investigate the modulation of these five mitonuclear associations by disease status (**Supplementary Materials**) using data from unaffected and affected individuals. The interaction of disease and nSNP for all five combinations achieved p-values below 6.0×10^−5^, suggesting that the association between mtSNPs and nSNPs is influenced by disease status (**Table 1**). Usually, interest focuses on the effect of mitonuclear combinations on disease risk, i.e., the effect of the interaction between mtSNP and nSNP on disease status [24]. Thus, we investigated the interaction terms for the five mitonuclear combinations (**Supplementary Materials**). They all achieved p-values <7.0×10^−5^ and odds ratios ≥ 1.5, suggesting that the combination of mtSNP and nSNP alleles modify disease risk (**Table 1**).

## Discussion

We began this study by using a single cohort approach to detect mitonuclear associations. We were able to identify such associations in a disease cohort but not amongst a control population, which may reflect the limited size of the cohort investigated. The results of previously published analyses are contradictory and do not always take population structure into account [20-22]. Thus, a strength of the approach we adopted is that it allows us to account for population stratification using covariates.

The lack of significant associations among the unaffected cohort contrasts with the results we obtained for the PD cohort, where five mitonuclear combinations showed a significant association. These combinations were also associated with PD risk.

Two mtSNPs, mt.2706 and mt.11467, were involved in significant combinations and both define haplogroups (H and U respectively [23]) that are associated with PD-risk [5, 12]. However, in our data, these mtSNPs do not achieve a significant association with PD on their own (**Table 1**). The nuclear components of these combinations map to three different locations. The nuclear component of rs1606610-mt.11467, maps to the intergenic region, downstream of *AIPL1* (**SFigure 5**). However, the paucity of supporting SNPs in LD means this result should be interpreted with caution. For rs11666267-mt.2706, the nuclear variant maps to *NIBAN3* (**SFigure 3**). ClinVar includes a report associating this rare germline variant with cerebrovascular disease (ClinVar ID:SCV001338733). Further, the polymorphism is adjacent to the 5’ UTR of *GLT25D1*, which encodes a collagen modifying enzyme, and an association with cerebrovascular disease has been reported [24]. Cerebrovascular disease has been postulated to be a PD-risk factor [25].

The nuclear components of the remaining significant combinations (all with mt.11467) map to *TBCA* (**SFigure 4**). *TBCA* encodes a tubulin chaperone and SNPs in the gene have been reportedly associated with PD age of onset [26]. In addition, its expression is altered in PD patients with dementia [27] and is elevated in the substantia nigra of PD patients [28]. In our study, the nuclear components of combinations mapping to TBCA achieved p-values between 10^−3^ and 6×10^−3^ (**Table 1**) in the association with PD, with odds ratios around 1.1.

It should also be noted that in a previous publication we identified several nSNPs showing associations with PD in the cohorts used here [29, 30] (**SFigure 6)**. The p-values for the interactions of any of the mitonuclear combinations involving the lead SNPs (chr4 rs2736990, chr6 rs3094609 and chr17 rs393152) are all above 0.05.

Although our analyses are adequately powered to detect relatively large effects, our results should be replicated in a larger cohort for validation and to enable subtler associations to be detected. Nonetheless, we present an approach that allows us to identify associations between mitochondrial and nuclear variants whilst accounting for population stratification and show that mitonuclear combinations can contribute to Parkinson’s disease risk.

## Data Availability

All data produced in the present study are available upon reasonable request to the authors

## Authors’ contributions

All authors contributed to acquisition and interpretation of data and approved the final manuscript. SP, GH and MSK performed the data analysis. GH and MSK designed the studies and supervised the work and drafted the manuscript.

## Acknowledgements

GH and SP are supported by the Wellcome Centre for Mitochondrial Research (203105/Z/16/Z).

## Supplementary Materials

### A) Materials and Methods Genotyping data and quality control

Welcome Trust Case/Control Consortium 2 (WTCCC2, https://www.wtccc.org.uk/ccc2/) single nucleotide polymorphism (SNP) genotyping data were downloaded from the European Genome Phenome Archive (ega-archive.org, 1958 British Birth Cohort, 58C=EGAD00000000021, n=2785; National Blood Donors, NBS=EGAD00000000023, n=2590 and Parkinson’s disease, PD=EGAS00000000034, n=2210). In brief, for all datasets, we excluded data from samples that did not meet the following criteria: minimum call rate >99% per sample, concordance between reported sex and estimated sex from X-chromosome-heterogeneity and removal of 1st-degree relatives (pi_hat threshold of <0.188). SNP exclusion criteria were; the proportion of missing calls >1%, Hardy Weinberg equilibrium p values <10^−7^ and minor allele frequencies (MAFs) <1%. Similar to previous work [32], ancestry was assessed using the 1000G v3 genotyping data (https://www.internationalgenome.org/data) through autosomal principal components and samples of non-European ancestry were removed. All these analyses were performed using PLINK v1.9 [33, 34]. The data from the 1958 Birth Cohort and the National Blood Service Donors, i.e. the two sets of unaffected individuals, were combined and herein referred to as 58C/NBS. Final datasets are summarised in **STable 1**.

#### Statistical Power

Statistical power was assessed using Quanto v1.2.4 [31]. Our power to detect mitocnuclear associations under an additive model, assuming a nDNA minor allele frequency of >10%, a mtDNA minor allele frequency >20%, an alpha of 5.0×10^−8^, an odds ratio greater than 1.5 and a population size >2100, was >0.80.

#### Selection of mtSNPs

Pairwise associations between all mtSNPs included in the genotyping data were assessed for SNPs with a minor allele frequency greater than 1% using HapMap Phase 3 European sequence data (https://www.sanger.ac.uk/resources/downloads/human/hapmap3.html, [36]) and PLINK, v1.9 [33, 34] (**STable 2**). This allowed us to select a set of mtSNPs where the call rate was high (>0.99) and the R^2^ between any pair mtSNPs in the set was below 0.8 (see results).

#### Association analyses

Logistic regression was carried out using Plink, v1.9 [33, 34] with sex and, to account for population stratification, the first 10 principal components of the autosomal SNPs as covariates. Association results were visualised using LocusZoom [32].

#### Mitonuclear association

To assess associations between nDNA and mtDNA variants one GWAS was carried out for each of the mtSNPs examined for each cohort using the presence of the variant mtDNA SNP allele as the outcome. To account for multiple testing, false discovery rates (FDR) were calculated across all nDNA and mtDNA SNPs using p.adjust in R v3.6.2 [37].

To investigate the modulation of mitonuclear associations by disease status for selected mitonuclear combinations, the cohorts of affected and unaffected, i.e. PD and 58C/NBS, were combined, the disease status included as a covariate and the significance of the nSNP/disease status interaction assessed. Analysis was carried as described above, i.e. using PLINK and including sex and 10 principal components derived from the autosomal SNPs. Since only five combinations were included we report here only the nominal p values.

#### Disease association

The association between DNA variants (mt and nSNPs) and disease status was carried out as described above using the combined data (i.e. from cohorts PD and 58C/NBS). To assess the effects of mitonuclear combinations, disease status was regressed onto the nuclear and mitochondrial genotypes and the significance of the interaction term was assessed. The presence of the mitochondrial variant was included as a covariate in the computations. These were otherwise performed as described above.

### B) Supplementary Data

Supplementary Tables 1-3

Supplementary Figures 1-6

## References

1. Chinnery, P.F. and G. Hudson, Mitochondrial genetics. Br Med Bull, 2013. 106: p. 135–59.

2. Lopez Sanchez, M.I.G., et al., Establishing risk of vision loss in Leber hereditary optic neuropathy. Am J Hum Genet, 2021. 108(11): p. 2159–2170.

3. Pickett, S.J., et al., Phenotypic heterogeneity in m.3243A>G mitochondrial disease: The role of nuclear factors. Ann Clin Transl Neurol, 2018. 5(3): p. 333–345.

4. Hudson, G., et al., Identification of an X-chromosomal locus and haplotype modulating the phenotype of a mitochondrial DNA disorder. Am J Hum Genet, 2005. 77(6): p. 1086–91.

5. Hudson, G., et al., Two-stage association study and meta-analysis of mitochondrial DNA variants in Parkinson disease. Neurology, 2013. 80(22): p. 2042–8.

6. Hudson, G., et al., Recent mitochondrial DNA mutations increase the risk of developing common late-onset human diseases. PLoS Genet, 2014. 10(5): p. e1004369.

7. Ghezzi, D., et al., Mitochondrial DNA haplogroup K is associated with a lower risk of Parkinson’s disease in Italians. Eur J Hum Genet, 2005. 13(6): p. 748–52.

8. Tranah, G.J., et al., Mitochondrial DNA sequence variation in multiple sclerosis. Neurology, 2015. 85(4): p. 325–30.

9. Vyshkina, T., et al., Genetic variants of Complex I in multiple sclerosis. J Neurol Sci, 2005. 228(1): p. 55–64.

10. Rollins, B., et al., Mitochondrial variants in schizophrenia, bipolar disorder, and major depressive disorder. PLoS One, 2009. 4(3): p. e4913.

11. Goncalves, V.F., et al., Examining the role of common and rare mitochondrial variants in schizophrenia. PLoS One, 2018. 13(1): p. e0191153.

12. Marom, S., M. Friger, and D. Mishmar, MtDNA meta-analysis reveals both phenotype specificity and allele heterogeneity: a model for differential association. Sci Rep, 2017. 7: p. 43449.

13. Samuels, D.C., et al., The power to detect disease associations with mitochondrial DNA haplogroups. Am J Hum Genet, 2006. 78(4): p. 713–20.

14. Kang, E., et al., Mitochondrial replacement in human oocytes carrying pathogenic mitochondrial DNA mutations. Nature, 2016. 540(7632): p. 270–275.

15. Yamada, M., et al., Genetic Drift Can Compromise Mitochondrial Replacement by Nuclear Transfer in Human Oocytes. Cell Stem Cell, 2016. 18(6): p. 749–754.

16. Hyslop, L.A., et al., Towards clinical application of pronuclear transfer to prevent mitochondrial DNA disease. Nature, 2016. 534(7607): p. 383–6.

17. Kang, E.J., et al., Mitochondrial replacement in human oocytes carrying pathogenic mitochondrial DNA mutations. Nature, 2016. 540(7632): p. 270-+.

18. Wei, W., et al., Germline selection shapes human mitochondrial DNA diversity. Science, 2019. 364(6442).

19. Kivisild, T., Maternal ancestry and population history from whole mitochondrial genomes. Investig Genet, 2015. 6: p. 3.

20. Sloan, D.B., P.D. Fields, and J.C. Havird, Mitonuclear linkage disequilibrium in human populations. Proc Biol Sci, 2015. 282(1815).

21. Yamamoto, K., et al., Genetic and phenotypic landscape of the mitochondrial genome in the Japanese population. Commun Biol, 2020. 3(1): p. 104.

22. Yonova-Doing, E., et al., An atlas of mitochondrial DNA genotype-phenotype associations in the UK Biobank. Nat Genet, 2021. 53(7): p. 982–993.

23. van Oven, M. and M. Kayser, Updated comprehensive phylogenetic tree of global human mitochondrial DNA variation. Hum Mutat, 2009. 30(2): p. E386–94.

24. Miyatake, S., et al., Biallelic COLGALT1 variants are associated with cerebral small vessel disease. Ann Neurol, 2018. 84(6): p. 843–853.

25. Kummer, B.R., et al., Associations between cerebrovascular risk factors and parkinson disease. Ann Neurol, 2019. 86(4): p. 572–581.

26. Tirozzi, A., et al., Genomic Overlap between Platelet Parameters Variability and Age at Onset of Parkinson Disease. Applied Sciences, 2021. 11(15): p. 6927.

27. Stamper, C., et al., Neuronal gene expression correlates of Parkinson’s disease with dementia. Mov Disord, 2008. 23(11): p. 1588–95.

28. Werner, C.J., et al., Proteome analysis of human substantia nigra in Parkinson’s disease. Proteome Sci, 2008. 6: p. 8.

29. Consortium, U.K.P.s.D., et al., Dissection of the genetics of Parkinson’s disease identifies an additional association 5’ of SNCA and multiple associated haplotypes at 17q21. Hum Mol Genet, 2011. 20(2): p. 345–53.

30. Hamza, T.H., et al., Common genetic variation in the HLA region is associated with late-onset sporadic Parkinson’s disease. Nat Genet, 2010. 42(9): p. 781–5.

31. Gauderman, W.J., Sample size requirements for matched case-control studies of gene-environment interaction. Stat Med, 2002. 21(1): p. 35–50.

32. Pruim, R.J., et al., LocusZoom: regional visualization of genome-wide association scan results. Bioinformatics, 2010. 26(18): p. 2336–2337.

