## Supplementary Figures for "Interactions between nuclear and mitochondrial SNPs and Parkinson’s disease risk"

### *Index*

**SFigure 1 | QQ plots of mitonuclear associations in 58C/NBS combined.**

**SFigure 2 | QQ plots of mitonuclear associations in PD**

**SFigure 3 | LocusZoom plot of the region containing rs11666267.**

**SFigure 4 | LocusZoom plot of the region containing rs254412, rs384109 and rs441492.**

**SFigure 5 | LocusZoom plot of the region containing rs1606610.**

**SFigure 6 | SFigure 10 | QQ plot of PD versus controls.**

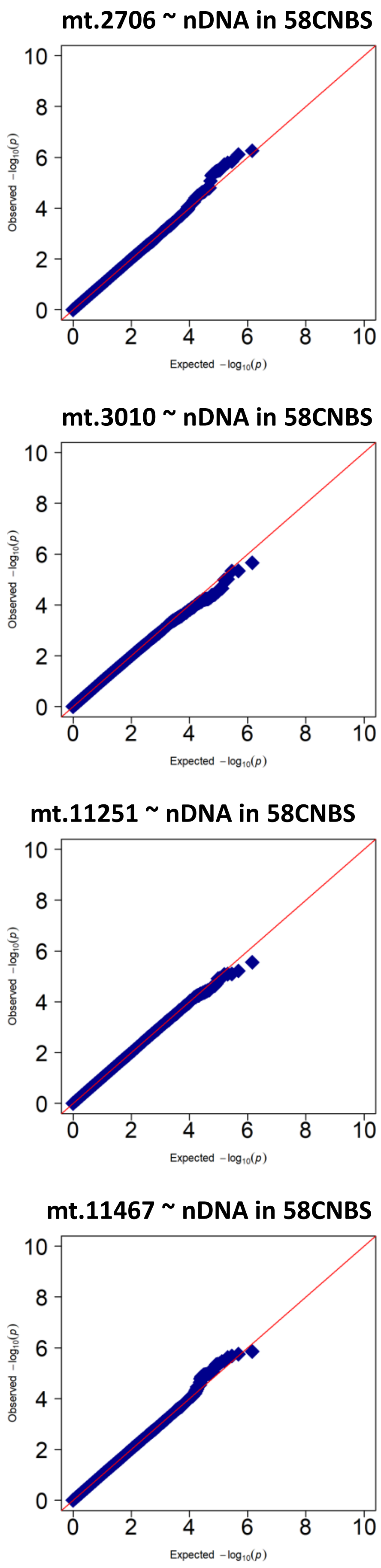


**SFigure 1 | QQ plots of mitonuclear associations in 58C/NBS cohorts combined.**
QQplots are shown for each of the selected mtSNPs (mt.2706, mt.3010, mt.11251 and mt.11467) in combined as 58C/NBS. Note, no nSNP reached achieved a P-value of <5x10-8.

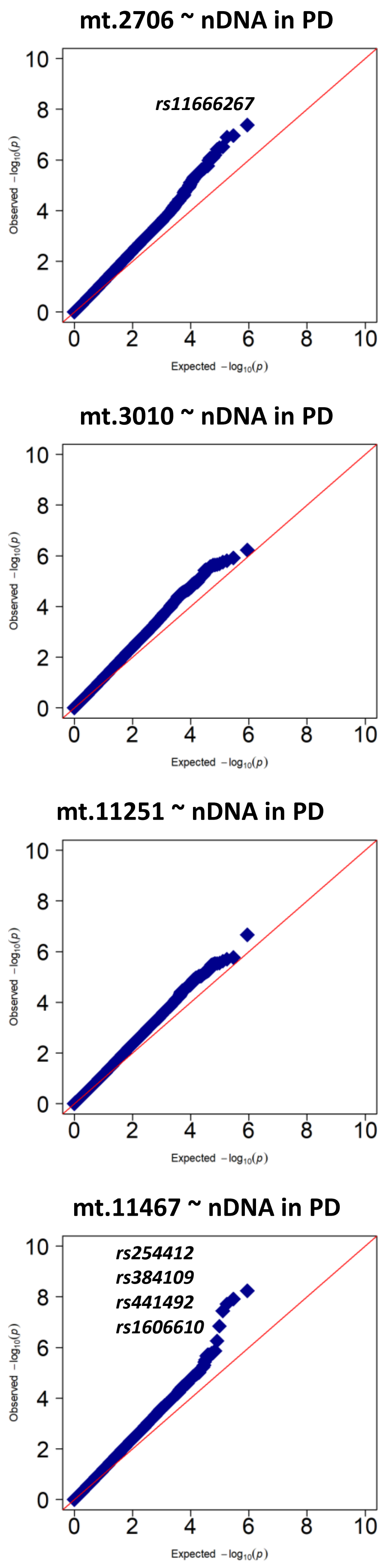


**SFigure 2 | QQ plots of mitonuclear associations in PD**

QQplots are shown for each of the selected mtSNPs (mt.2706, mt.3010, mt.11251 and mt.11467) in PD. Note,  nSNPs achieving a P-value of <5x10-8 are highlighted.

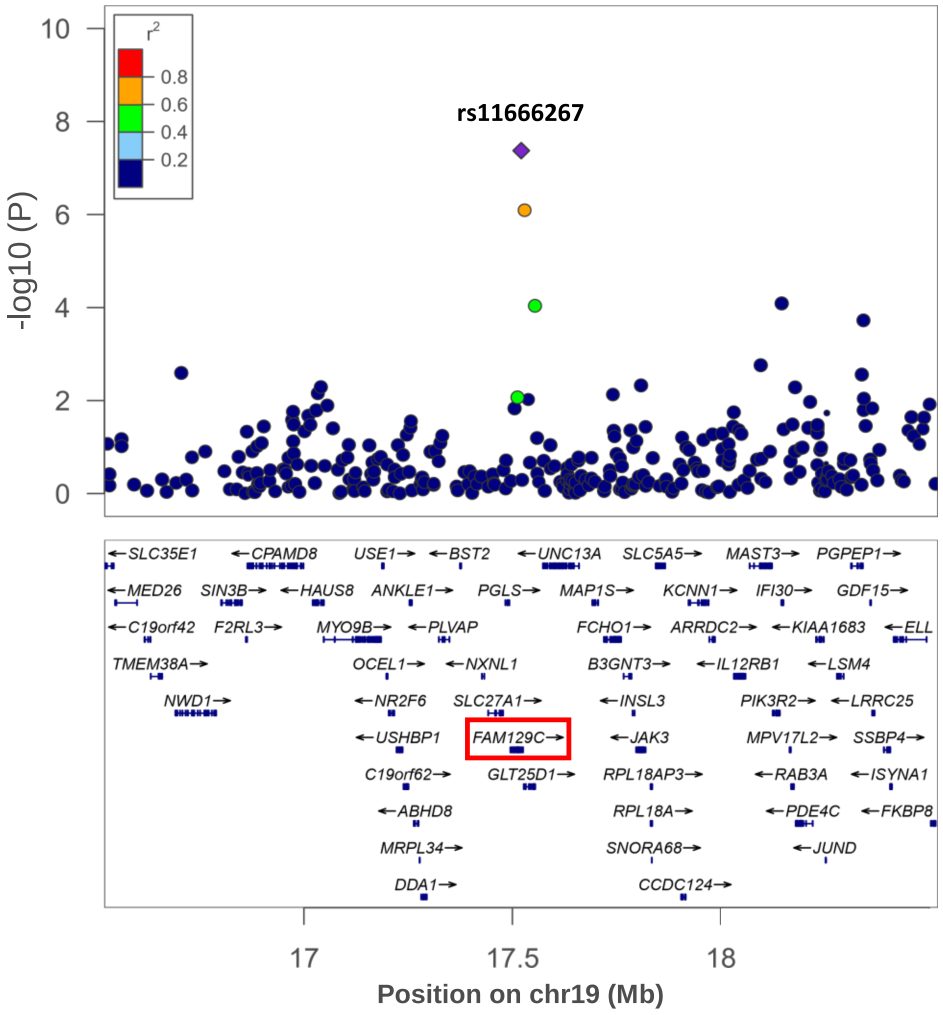


**SFigure 3 | LocusZoom plot of the region containing rs11666267.**

LocusZoom plot of the region (+/- 1000KB) surrounding rs11666267 (**Table 1**). $R^2$ determined using HapMap CEU samples. FAM129C is also known as NIBAN3.

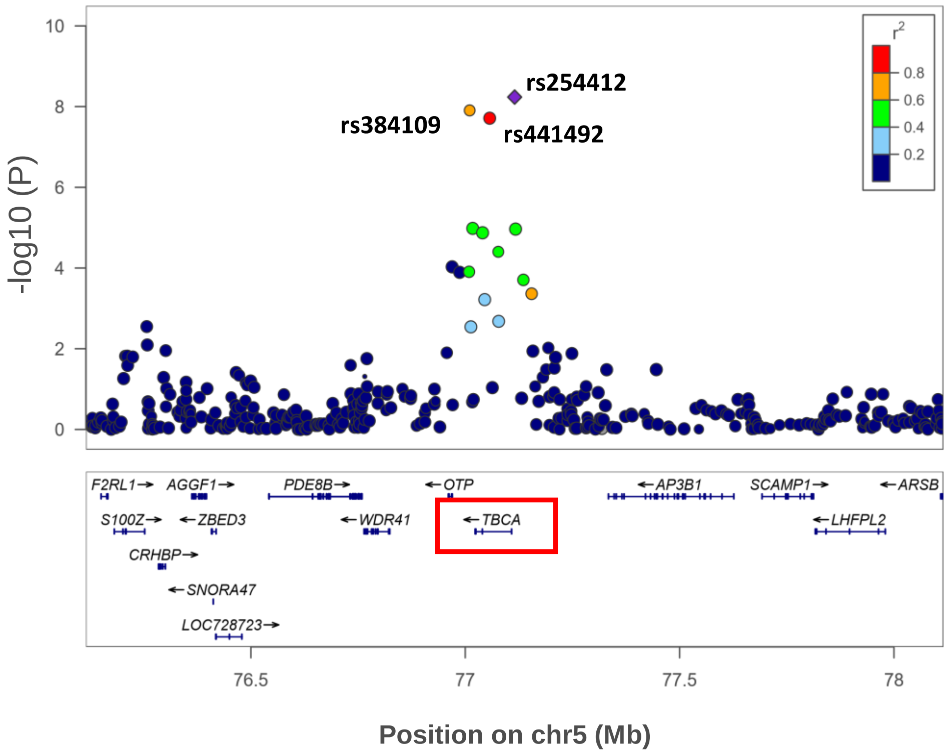


**SFigure 4 | LocusZoom plot of the region containing rs254412, rs384109 and rs441492.**

LocusZoom plot of the region (+/- 1000KB) surrounding rs254412, rs384109 and rs441492 (**Table 1**). $R^2$ determined using HapMap CEU samples.

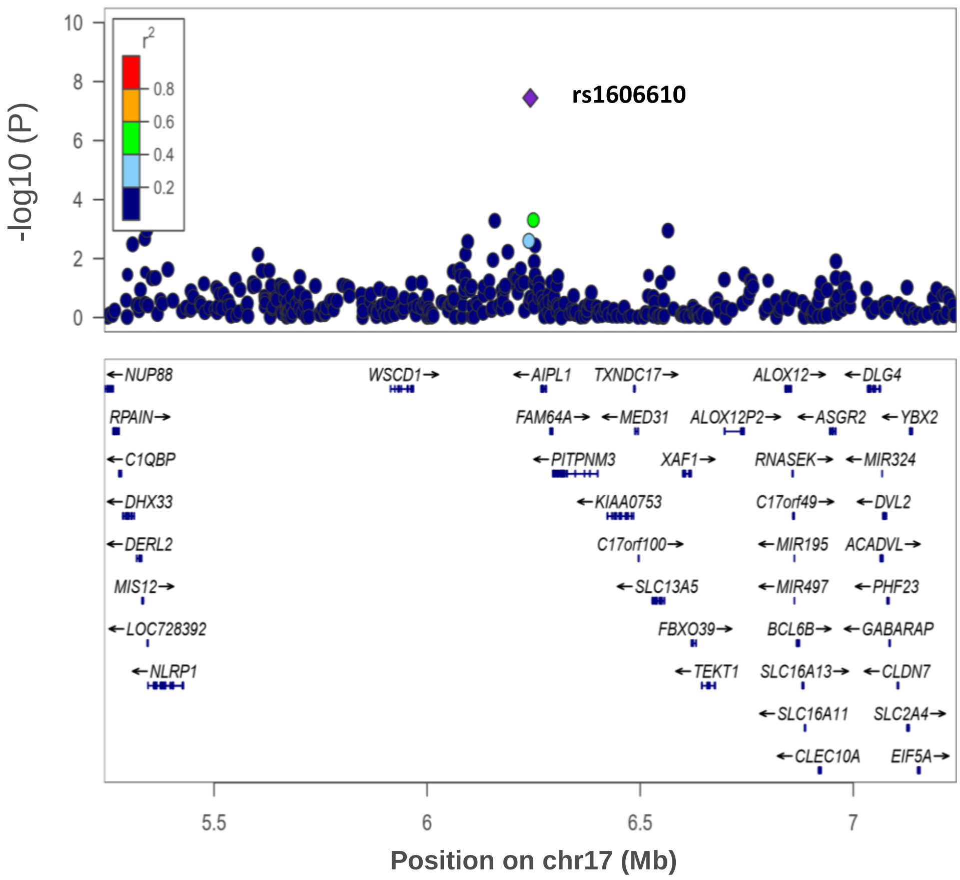


**SFigure 5 | LocusZoom plot of the region containing rs1606610.**

LocusZoom plot of the region (+/- 1000KB) surrounding rs1606610 (Table 1). $R^2$ determined using HapMap CEU samples.

a)

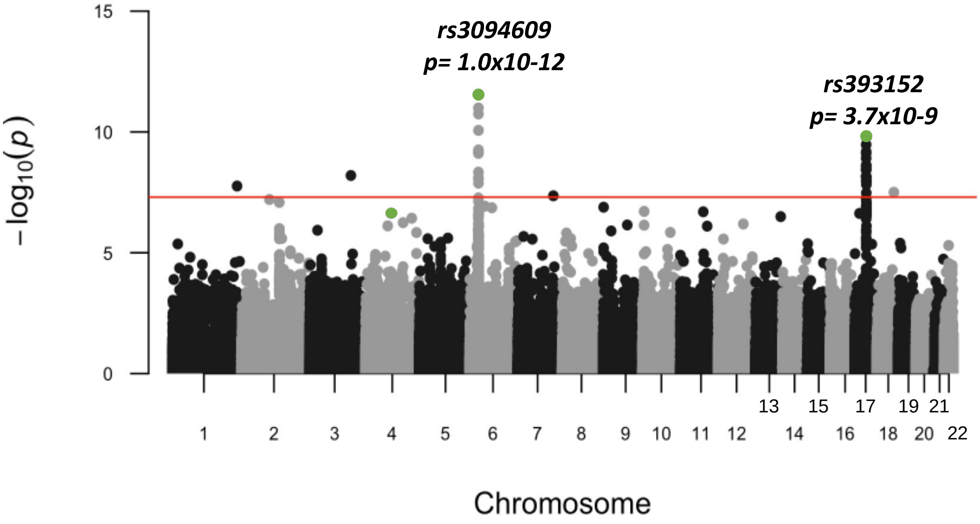


a)

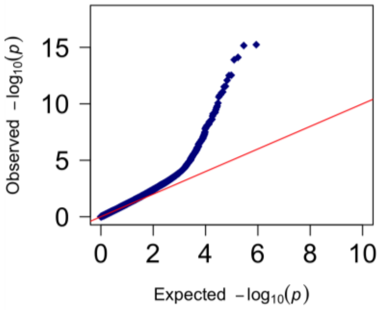


**SFigure 6 | nSNPs in PD versus Controls (58CNBS).**

Manhattan plot (a) and corresponding Q-Q plot (b) of the association between nSNPs in PD cases versus controls (58CNBS). The Q-Q plot shows significant (5.0x10$^{-8}$) association between PD and 55 nSNPs, which is inline with previous observations.
